# Assessing the Competing Risks of Loss to Follow-up and Mortality Among Adults Receiving Antiretroviral Therapy in Northern Ghana: Application of the Fine-Gray Subdistribution Hazard Model

**DOI:** 10.64898/2026.10.15.26365612

**Authors:** Mohammed Yushawu Abdulai, Solomon Crusader Abo, Benjamin Tommy Bavug

## Abstract

**Background:** Retention in antiretroviral therapy (ART) programmes remains a major public health challenge in HIV care, with loss to follow-up (LTFU) and death threatening treatment success. Because these outcomes are competing events, conventional survival methods may yield biased estimates of their incidence and associated risk factors.

**Objective:** To estimate the cumulative incidence of LTFU and mortality and identify factors associated with these outcomes among adults receiving ART in Northern Ghana using a competing-risks framework.

**Methods:** We conducted a retrospective cohort study of 329 adults living with HIV who initiated ART at Nalerigu Baptist Medical Centre, Ghana, between January 2013 and December 2019. Fine–Gray subdistribution hazard models were used to evaluate predictors of LTFU and death while accounting for competing risks. Cause-specific Cox proportional hazards models were fitted as a sensitivity analysis.

**Results:** The incidence rates were 2.2 per 100 person-months (95% CI: 1.9–2.6) for LTFU and 0.74 per 100 person-months (95% CI: 0.57–0.98) for death. At 36 months, the cumulative incidence was 50.2% for LTFU and 16.0% for death. Patients residing closer to the clinic had a lower cumulative incidence of LTFU than those living farther away (SHR = 0.68, 95% CI: 0.49–0.93), whereas higher baseline weight was associated with increased LTFU (SHR = 1.02 per kg, 95% CI: 1.00–1.04). Mortality was lower among patients receiving efavirenz-based regimens (SHR = 0.27, 95% CI: 0.15–0.50), females (SHR = 0.39, 95% CI: 0.18–0.85), and those with higher baseline weight (SHR = 0.93 per kg, 95% CI: 0.90–0.96). Findings were broadly consistent in cause-specific Cox models.

**Conclusion:** LTFU was substantially more frequent than mortality among adults receiving ART in Northern Ghana. Travel distance to the treatment facility was an important barrier to retention, while ART regimen, sex, and baseline weight were associated with mortality. Expanding access to ART services through decentralized and community-based models may improve retention and support efforts to achieve HIV control targets.

## 1 Introduction

Retention in antiretroviral therapy (ART) programmes remains a major challenge in the global HIV/AIDS response, with attrition driven largely by loss to follow-up (LTFU) and mortality. Sustained engagement in care is essential to achieve viral suppression, reduce onward transmission, and improve survival [8, 26–28].

Globally, an estimated 40.8 million people were living with HIV at the end of 2024, with approximately 1.3 million new infections and about 630,000 AIDS-related deaths occurring that year; women and girls accounted for 53% of all people living with HIV worldwide [23]. Sub-Saharan Africa remains the region most heavily affected, with approximately 26.5 million people living with HIV in 2024, as about 65% of the global total, of whom 82% were receiving ART [23]. In Ghana, an estimated 337,435 people were living with HIV as of the most recent Ghana AIDS Commission report in 2019, with a treatment cascade of approximately 70-81-92 percent aware of status, on treatment, and virally suppressed, respectively. This progress remains below the UNAIDS 95-95-95 targets [12].

Programme retention varies widely across settings: high attrition has been reported in multiple African cohorts, with LTFU rates exceeding 10 per 100 person-years in some programmes [17, 18, 21]. When patients die, they can no longer become LTFU, and vice versa, making these competing risks. Standard Kaplan–Meier [14] and Cox proportional hazards [9] methods that censor competing events overestimate cumulative incidence. The Fine-Gray proportional subdistribution hazard model [11] addresses this by directly modelling covariate effects on the cumulative incidence function (CIF) in the presence of competing risks. Correct specification requires that covariate effects on the subdistribution hazard be proportional over time; the study formally assessed this assumption.

Loss to follow-up remains a major obstacle to the long-term success of antiretroviral therapy programmes, particularly in resource-limited settings where patient retention is essential for sustained viral suppression and improved survival. Although several studies have investigated predictors of loss to follow-up among patients receiving ART in sub-Saharan Africa, many have relied on conventional survival methods that treat competing events such as death as independent censoring events. Such approaches may produce biased estimates of event probabilities and covariate effects when competing risks are present. Furthermore, evidence from Northern Ghana using competing-risk methodology remains limited. Therefore, this study applies the Fine-Gray subdistribution hazard model to evaluate factors associated with loss to follow-up while appropriately accounting for the competing risk of death among HIV patients receiving antiretroviral therapy. Specifically, the study estimates cumulative incidence functions for loss to follow-up and mortality and identifies predictors associated with these outcomes.

This study quantifies the incidence and determinants of LTFU and death among adults initiating ART at Nalerigu Baptist Medical Centre in Northern Ghana (2013–2019), where no prior competing risks analysis has been reported.

## 2 Materials and Methods

This retrospective cohort study used longitudinal program data from the National AIDS/STI Control Programme (NACP), obtained from the Antiretroviral Therapy (ART) unit of the Nalerigu Baptist Medical Centre in the North East Region of Ghana. The study included records of adults aged ≥ 15 years who initiated ART between January 1, 2013, and December 31, 2019. Participants were followed from ART initiation until loss to follow-up (LTFU), death, transfer to another facility, or the end of the study period.

Participants were eligible if they: (i) were aged ≥ 15 years at ART initiation; (ii) initiated first-line ART at the study facility during the accrual period; and (iii) had at least one scheduled follow-up date or documented post-initiation contact that enabled outcome classification. Patients were excluded if they had missing or implausible ART initiation dates, irreconcilable duplicate patient identifiers, transferred into the clinic without a verifiable ART initiation date, or had no post-initiation clinical information.

This study was reported in accordance with the Strengthening the Reporting of Observational Studies in Epidemiology (STROBE) guidelines for observational cohort studies.

Baseline characteristics were measured at ART initiation. Sociodemographic variables included age in years, sex (male/female), marital status (single, married, divorced/separated, or widowed), educational status (any formal education versus none), employment status (employed versus not employed), and religious affiliation (Christian, Muslim, Traditional, or other). Travel distance to the clinic was categorized as short (≤ 10 km) or long (*>* 10 km). Clinical and treatment-related baseline variables included body weight in kilograms, the non-nucleoside reverse transcriptase inhibitor (NNRTI) component of the ART regimen (Efavirenz [EFV] versus Nevirapine [NVP]), hospital admission at ART initiation (yes/no), provision of a contact telephone number (yes/no), and calendar year of ART initiation. Year of ART initiation was included to account for potential programmatic changes over time.

The analysis considered two mutually exclusive time-to-event outcomes: LTFU and death. LTFU was defined as the absence of a clinic visit, pharmacy refill, or other documented clinical contact for ≥ 90 days beyond the scheduled appointment or medication-refill date. All-cause mortality was determined from clinical or inpatient records and confirmed through programme tracing. Individuals who initially met the LTFU definition but were subsequently found to have died before reaching the 90-day threshold were reclassified as deceased. Individuals who transferred to another facility before meeting the LTFU criterion were censored at the transfer date. Participants who remained alive and in care at the end of follow-up without LTFU or death were censored administratively at their last documented clinical contact or at the study end date, as applicable. Let *T_i_* denote the time from ART initiation to the first observed event for individual *i*, and let *ɛ_i_* ∈ {0, 1, 2} denote the event type, where 0 denotes censoring, 1 denotes LTFU, and 2 denotes death. Since death prevents subsequent LTFU and LTFU precludes observation of subsequent death within the study cohort, the outcomes were analysed as competing risks.

For event type *k* ∈ {1, 2}, the cumulative incidence function (CIF) was defined as

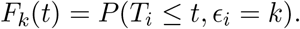

The CIF represents the probability that an individual experiences event type *k* by time *t*, while appropriately accounting for the occurrence of the competing event. The corresponding subdistribution hazard for event type *k* was defined as

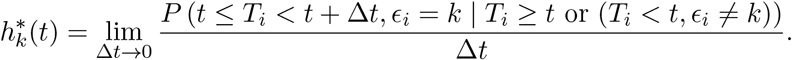

Unlike the cause-specific hazard, the Fine–Gray subdistribution hazard retains individuals who have experienced the competing event in the risk set using appropriate weighting. This feature enables direct modelling of covariate effects on the cumulative incidence of the event of interest.

The Fine–Gray subdistribution hazard model was selected because the primary objective was to assess the association between baseline covariates and the cumulative incidence of LTFU in the presence of death as a competing event. A second Fine–Gray model was fitted to evaluate the cumulative incidence of death in the presence of LTFU as the competing event. Unlike cause-specific hazard models, which model the instantaneous event rate among individuals who remain free of all events, the Fine–Gray model directly evaluates covariate effects on the CIF. This interpretation is particularly relevant for clinical and public-health planning, where the interest is commonly in the absolute probability of LTFU or death over time in the presence of competing outcomes.

For event type *k*, the Fine–Gray model was specified as

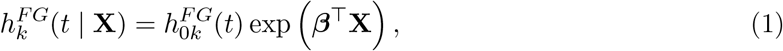

where *h_k_^FG^*(*t* | **X**) is the subdistribution hazard for event type *k* at time *t*, *h*_0_*_k_^FG^*(*t*) is the baseline subdistribution hazard, **X** is the vector of covariates, and ***β*** is the vector of regression coefficients. The exponentiated regression coefficients, exp(*β_j_*), are interpreted as subdistribution hazard ratios (SHRs). An SHR greater than one indicates a higher cumulative incidence of the outcome of interest over follow-up, conditional on the competing-risk structure. Separate Fine–Gray models were fitted for LTFU and death. In the LTFU model, death was specified as the competing event; in the mortality model, LTFU was specified as the competing event.

Covariate selection was guided by statistical and clinical considerations. Variables identified in previous studies of HIV retention, LTFU, and mortality as potential predictors were included in the multivariable models irrespective of their univariable statistical significance. This full-model approach was used to provide adjustment for clinically relevant potential confounders and to avoid the instability and selection bias that may arise from stepwise variable-selection procedures.

Data quality assessments were undertaken before statistical analysis. All study variables were screened for missing values and data inconsistencies. Following data verification, all variables included in the competing-risks models contained complete observations. Consequently, no participants were excluded because of missing covariate data, and all 329 eligible patients were retained in the analysis. Although no formal sample-size calculation was performed owing to the retrospective nature of the study, the number of observed mortality and loss-to-follow-up events was sufficient relative to the number of covariates included in the multivariable models, exceeding commonly recommended event-per-variable criteria for survival regression analyses.

As a sensitivity analysis, cause-specific Cox proportional hazards models were fitted for both outcomes. For the LTFU analysis, death was treated as a censoring event; for the mortality analysis, LTFU was treated as a censoring event. Estimates from the cause-specific Cox models were compared with those from the Fine–Gray models to assess the robustness of the identified risk factors and to distinguish associations with the event-specific hazard from associations with cumulative incidence.

All statistical analyses were performed using R version 4.3.1. The cmprsk package was used to estimate CIFs with the cuminc function and to fit Fine–Gray regression models using the crr function. The proportional subdistribution hazards assumption was assessed using the psh.test function from the crrSC package, supported by scaled Schoenfeld-type residual plots. Descriptive statistics were generated using SPSS. Additional simulation studies assessing finite-sample performance are presented in the Supplementary Materials.

## 3 Results

A total of 329 HIV-positive adults initiating ART between 2013 and 2019 were included in the study. Examination of the dataset revealed no missing values among the variables considered in the analysis; therefore, all 329 patients contributed to the competing-risks modelling. Table A1 presents the demographic and socio-economic characteristics of the study population. In a total of 329 respondents, 232 were female (70.5%) and 97 were male (29.5%). The majority of patients (53.8%) were between 30 and 49 years of age, followed by 35.3% aged 15–29 years, and 10.9% aged 50 years or older. Religious affiliation was predominantly Muslim (53.5%), with Christians accounting for 37.4% and traditionalists 9.1%. Marital status was varied: 17% were single, 66.3% married, 11.2% widowed, and 5.5% divorced or separated. As shown in Table A1, over half of the patients (56.5%) had no formal education, while the remaining 43.5% had attained at least primary education. Employment status was nearly evenly split, with 53.5% employed and 46.5% unemployed. Regarding access to care, 53.2% of patients resided within a short travel distance of the ART facility, while 46.8% travelled longer distances.

### 3.1 Assessment of the Proportional Subdistribution Hazard Assumption

Schoenfeld residual plots are diagnostic tools used to visually assess whether the proportional subdistribution hazards assumption holds for the Fine-Gray model. The proportional hazard assumption states that the effect of a covariate on the subdistribution hazard is constant over time[15]. To check this assumption, a scatterplot of the residuals against the failure time is created for the lost-to-follow-up and death subdistribution hazards model, and a smoother line is added to show the average trend. Figure 2 and Figure 3 display the Schoenfeld residual plots for the lost-to-follow-up and death subdistribution hazards models (Appendix). For both outcomes, the effects of the covariates on the subdistribution hazards appeared consistent over time, as the smoothed residual curves were approximately horizontal, visually confirming that the proportional subdistribution hazards assumption is met. Table 1 presents the test statistics and corresponding p-values for both LTFU and death as outcomes. None of the covariates showed statistically significant evidence of time-varying effects (p-value > 0.05), indicating that the proportional subdistribution hazards assumption is reasonably satisfied for all included variables. The global test also yielded non-significant results for both LTFU (p = 0.988) and death (p = 0.498), further supporting the adequacy of the proportionality assumption in this analysis, warranting the application of the Fine-Gray subdistribution hazard model for the ART dataset.

**Table 1:** Statistical Test for Assessing Proportional Hazard Assumption.

| Covariates | Lost to follow-up |  | Death |  |
| --- | --- | --- | --- | --- |
|  | Statistic | P-value | Statistic | P-value |
| Education | 0.8144 | 0.3668 | 1.1194 | 0.2900 |
| Age | 0.2467 | 0.6194 | 0.3953 | 0.5296 |
| Distance to facility | 1.8237 | 0.1769 | 2.6511 | 0.1035 |
| type of regimen at baseline | 0.3763 | 0.5396 | 1.7564 | 0.1851 |
| Employment | 2.7511 | 0.0972 | 2.6760 | 0.1019 |
| Gender | 1.0164 | 0.3134 | 0.0247 | 0.8751 |
| Marital: Divorced/Separated | 0.5225 | 0.4698 | 0.2418 | 0.6229 |
| Marital: Married | 0.2139 | 0.6438 | 0.1262 | 0.7224 |
| Marital: Widowed | 1.2239 | 0.2686 | 0.4023 | 0.5259 |
| Contact number | 2.1225 | 0.1451 | 0.0109 | 0.9167 |
| Weight | 0.4511 | 0.5018 | 0.1764 | 0.6745 |
| Hospital Admission | 0.0098 | 0.9211 | 0.6591 | 0.4169 |
| Overall Test | 2e-04 | 0.9876 | 0.4591 | 0.4981 |

### 3.2 Time-to-Event Analysis Using Cumulative Incidence Functions

Figure 1 displays the cumulative incidence functions for the competing events of loss to follow-up (LTFU) and death, stratified by gender. Overall, LTFU was the most common event across the study period. It is shown that females appeared to experience a higher cumulative incidence of LTFU descriptively; however, sex was not a statistically significant predictor in the multivariable Fine-Gray model. At 36 months, the cumulative incidence of LTFU was 50.1% for females and 46.2% for males. In contrast, the cumulative incidence of death was higher among males than females, reaching 25.5% and 13.9%, respectively, at the same time point.

**Figure 1:**
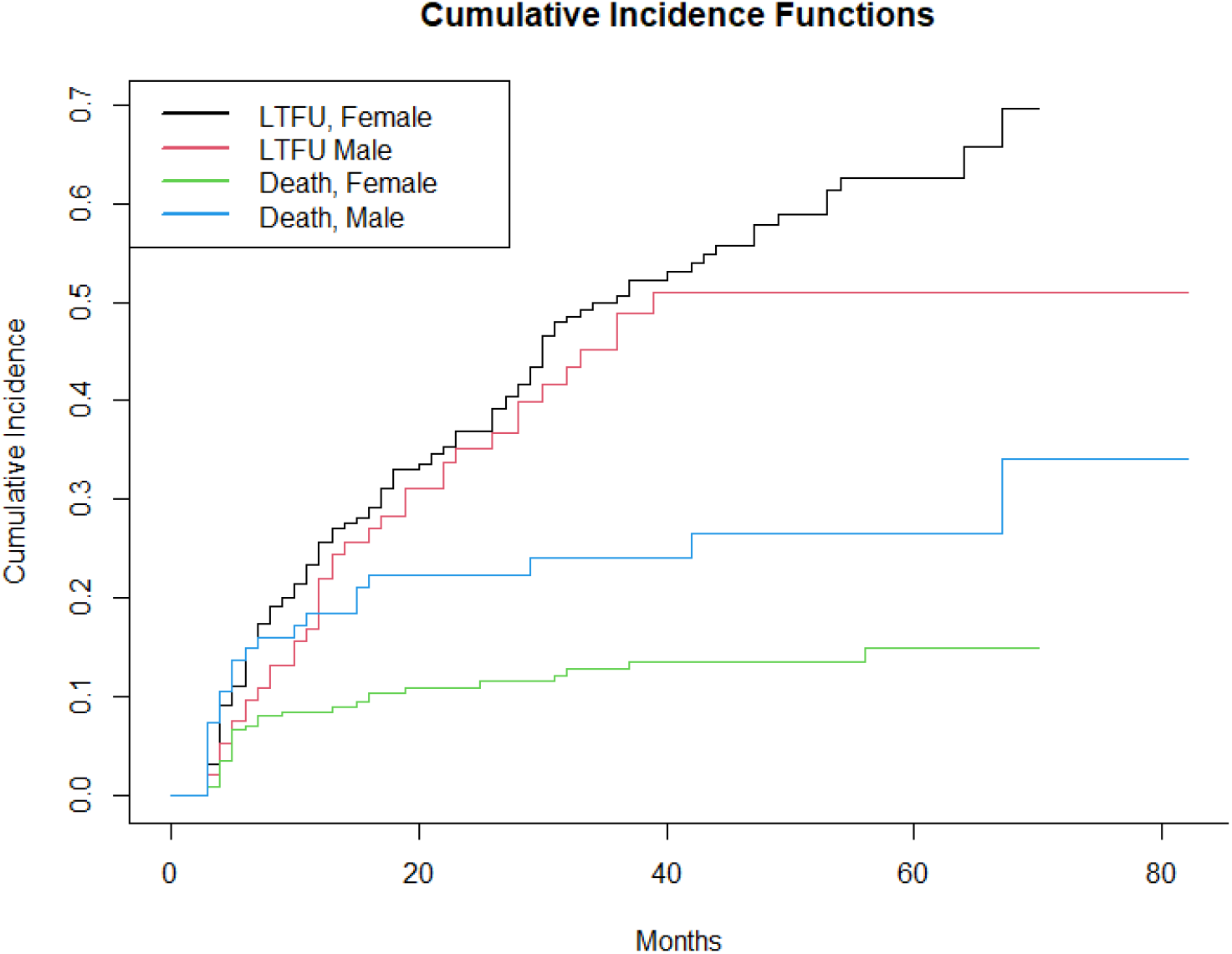
Cumulative incidence functions for LTFU and death, stratified by gender.

### 3.3 Incidence of LTFU and Death

Table 2 analyses the attrition among patients initiating antiretroviral treatment (ART), focusing on being lost to follow-up (LTFU) and death. The analysis covers 36 months and examines how various demographic and clinical factors influence the cumulative incidence of these outcomes. Table 2 presents the cumulative incidence of LTFU and death at 6, 12, 24, and 36 months, categorized by key patient characteristics such as sex, age, religion, and other factors.

**Table 2:**
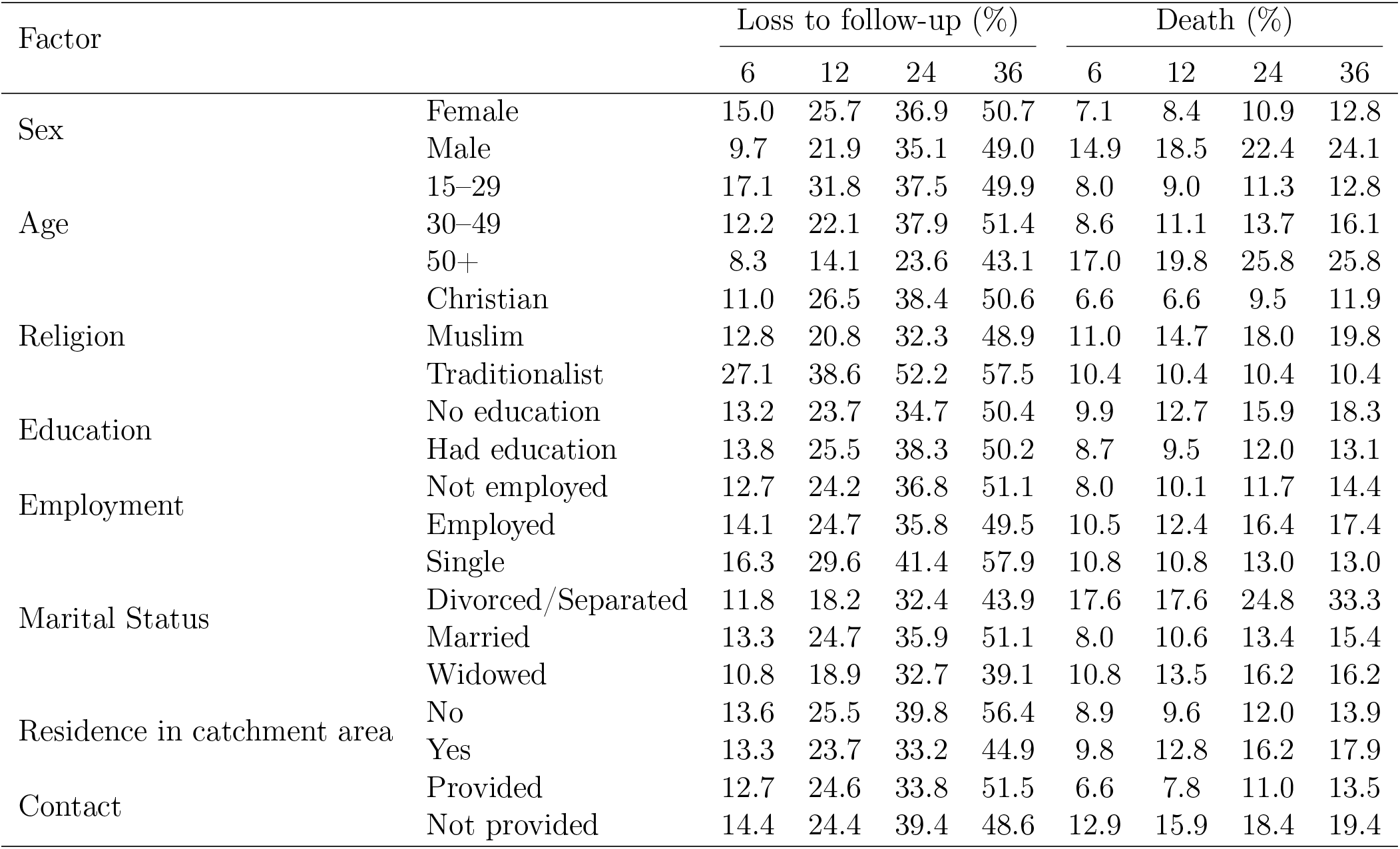
Cumulative Incidence of Attrition from HIV Care by Demographic Characteristics.

#### 3.3.1 Incidence of Loss to Follow-up

The incidence rate of LTFU was 2.2 per 100 person-months of observation (95% CI: 1.9–2.6), and 46.5% of patients were lost to follow-up during the study period. The cumulative incidence of LTFU increased steadily over time, reaching 13.4%, 24.5%, 36.3%, and 50.2% at 6, 12, 24, and 36 months, respectively. At 12 months, the cumulative incidence of LTFU was highest among younger patients aged 15–29 years (31.8%) and lowest among patients aged 50 years and above (14.1%). Higher LTFU was also observed among traditionalists (38.6%) and single individuals (29.6%). Differences according to sex, education, employment status, residence, and contact availability were relatively small. Detailed cumulative incidence estimates for all subgroups are presented in Table 2.

#### 3.3.2 Incidence of Death

The incidence rate of death was 0.74 per 100 person-months of observation (95% CI: 0.57–0.98), and 15.8% of patients died during follow-up. The cumulative incidence of death increased from 9.4% at 6 months to 16.0% at 36 months. At 12 months, mortality was higher among males (18.5%) than females (8.4%) and increased with age, reaching 19.8% among patients aged 50 years and above. Mortality was also relatively higher among Muslims (14.7%) and divorced/separated individuals (17.6%). A notable difference was observed according to ART regimen, with patients receiving NVP-based therapy experiencing a higher cumulative incidence of death than those receiving EFV-based therapy (28.4% vs. 5.5% at 12 months). Detailed estimates for all demographic and clinical subgroups are presented in Table 2.

### 3.4 Subdistribution Hazard Models for LTFU and Death

Table 3 presents the hazard ratios and 95% confidence intervals obtained from Fine-Gray subdistribution hazard models for both loss to follow-up (LTFU) and death. The models assess the association between demographic, clinical, and treatment-related covariates and the subdistribution hazard of each outcome, accounting for the presence of competing risks.

**Table 3:** Hazard Ratios from Fine-Gray Subdistribution Hazard Models for LTFU and Death.

| Covariate | LTFU |  |  | Death |  |  |
| --- | --- | --- | --- | --- | --- | --- |
|  | SHR | 95% CI | p-value | SHR | 95% CI | p-value |
| Education: No formal education | 0.960 | 0.679–1.356 | 0.818 | 0.855 | 0.471–1.552 | 0.605 |
| Age (per year) | 0.991 | 0.972–1.009 | 0.337 | 1.012 | 0.990–1.034 | 0.282 |
| Distance to facility: Short | 0.677 | 0.490–0.937 | 0.019 | 1.793 | 0.992–3.240 | 0.054 |
| Regimen: EFV-based | 1.167 | 0.810–1.681 | 0.403 | 0.272 | 0.148–0.500 | < 0.001 |
| Employment: Employed | 0.921 | 0.623–1.363 | 0.682 | 0.933 | 0.500–1.740 | 0.828 |
| Gender: Female | 1.237 | 0.767–1.994 | 0.382 | 0.393 | 0.182–0.850 | 0.018 |
| Marital status (ref: Single) |  |  |  |  |  |  |
| Divorced/Separated | 0.806 | 0.349–1.861 | 0.614 | 1.410 | 0.361–5.508 | 0.621 |
| Married | 0.862 | 0.556–1.336 | 0.506 | 1.420 | 0.561–3.591 | 0.459 |
| Widowed | 0.598 | 0.310–1.155 | 0.125 | 1.713 | 0.472–6.217 | 0.417 |
| Contact: Provided | 0.891 | 0.632–1.258 | 0.511 | 0.873 | 0.424–1.796 | 0.712 |
| Weight (per kg) | 1.022 | 1.004–1.041 | 0.018 | 0.932 | 0.903–0.963 | < 0.001 |
| Hospital admission: Admitted | 1.398 | 0.930–2.103 | 0.107 | 0.695 | 0.281–1.720 | 0.431 |

Note: SHR = subdistribution hazard ratio; CI = confidence interval; EFV = efavirenz.
Reference categories: Long distance, NVP-based regimen, Not employed, Male, Single, Contact not provided, Not admitted.

#### 3.4.1 Fine-Gray Subdistribution Hazard model for LTFU

From Table 3, only two covariates, namely distance to facility and baseline weight, were significantly associated with the cumulative incidence of LTFU (*p <* 0.05). Patients who travelled short distances to the treatment facility had a 32.3% lower subdistribution hazard of LTFU than those who travelled long distances (SHR = 0.677, 95% CI: 0.490–0.937, *p* = 0.019). This indicates that the cumulative incidence of LTFU was lower among patients residing closer to the treatment facility. Additionally, baseline weight was positively associated with LTFU. Specifically, a one-kilogram increase in baseline weight was associated with a 2.2% increase in the subdistribution hazard of LTFU (SHR = 1.022, 95% CI: 1.004–1.041, *p* = 0.018). Thus, higher baseline weight was associated with an increased cumulative incidence of LTFU.

#### 3.4.2 Fine-Gray Subdistribution Hazard Model for Death

In this model, death is treated as the event of interest, with LTFU as the competing event. Only three covariates (Type of NNRTI, Sex, and Weight) are statistically significant. Patients receiving EFV-based regimens experienced a lower cumulative incidence of mortality (72%); however, because treatment allocation was not randomized, residual confounding cannot be excluded. It also means that the cumulative incidence of death was lower for patients with the EFV-based regimen than for patients with the NVP-based regimen. Female patients had a lower SHR of death than that of the male category; thus, female sex was associated with a 60.7% lower subdistribution hazard of death relative to males. This also suggests that the cumulative incidence of death was lower for females than for males. Also, a 1-unit increase in a patient’s weight is associated with a 6.8% decrease in the SHR. It can also be inferred that a unit increase in a patient’s weight results in a decrease in the cumulative incidence of death.

### 3.5 Sensitivity Analysis

To evaluate the robustness of the Fine-Gray competing risks results, cause-specific Cox proportional hazards models were fitted for both LTFU and death. While Fine-Gray models estimate effects on cumulative incidence, cause-specific Cox models estimate effects on the instantaneous hazard among individuals still at risk. Consistency between the two approaches was used to assess robustness. Before fitting the cause-specific models, proportional hazards (PH) assumptions were assessed using Schoenfeld residual tests. Table 4 reports the Schoenfeld residual test statistics and corresponding p-values for each covariate and the global test, separately for the loss to follow-up (LTFU) and death cause-specific Cox proportional hazards models. No covariate violated the PH assumption at the 5% significance level in either model. Furthermore, the global Schoenfeld residual tests were not statistically significant for either the LTFU model (*χ*^2^ = 8.04, *df* = 10, *p* = 0.624) or the death model (*χ*^2^ = 8.09, *df* = 10, *p* = 0.620), indicating that the proportional hazards assumption was adequately satisfied throughout the follow-up period for both cause-specific models.

**Table 4:** Schoenfeld residual tests for the proportional hazards assumption in the cause-specific Cox models.

| Covariate | LTFU |  | Death |  |
| --- | --- | --- | --- | --- |
| | $\chi^2$ | *p*-value | $\chi^2$ | *p*-value |
| Education: No formal education | 0.013 | 0.908 | 1.508 | 0.219 |
| Age | 0.000 | 0.983 | 1.001 | 0.317 |
| Distance: Long | 2.350 | 0.125 | 0.082 | 0.775 |
| Regimen: NVP-based | 0.063 | 0.802 | 0.254 | 0.615 |
| Employment: Not employed | 1.646 | 0.199 | 3.742 | 0.053 |
| Gender: Male | 0.164 | 0.685 | 0.130 | 0.718 |
| Marital status: Single | 0.180 | 0.671 | 1.396 | 0.237 |
| Contact: Not provided | 0.460 | 0.498 | 0.006 | 0.939 |
| Weight | 0.207 | 0.649 | 0.100 | 0.752 |
| Hospital admission: No | 0.156 | 0.693 | 0.008 | 0.928 |
| **Global test** | **8.039** | **0.624** | **8.091** | **0.620** |

#### 3.5.1 Cause-Specific Sensitivity Analysis for LTFU

Table 5 presents the cause-specific Cox model estimates alongside the Fine-Gray subdistribution hazard model estimates for loss to follow-up (LTFU). The cause-specific model identified 153 LTFU events among 329 patients (concordance *C* = 0.568; likelihood ratio test: *χ*^2^ = 10.87, *df* = 10, *p* = 0.40). Overall, the results were broadly concordant with those from the Fine-Gray model.

**Table 5:** Performance comparison of Fine-Gray SHR and CS-HR for LTFU.

| Covariate | Fine-Gray SHR |  |  | Cause-Specific HR |  |  |
| --- | --- | --- | --- | --- | --- | --- |
| | Est. | 95% CI | $p$ | Est. | 95% CI | $p$ |
| Education: No formal education | 1.032 | 0.730–1.459 | 0.860 | 1.102 | 0.771–1.573 | 0.595 |
| Age (per year) | 0.989 | 0.971–1.007 | 0.220 | 0.992 | 0.975–1.010 | 0.394 |
| Distance: Long | 1.493 | 1.079–2.064 | 0.015 | 1.454 | 1.030–2.053 | 0.033 |
| Regimen: NVP-based | 0.865 | 0.601–1.246 | 0.440 | 1.268 | 0.861–1.868 | 0.230 |
| Employment: Not employed | 1.119 | 0.763–1.641 | 0.570 | 1.118 | 0.763–1.638 | 0.567 |
| Gender: Male | 0.859 | 0.542–1.360 | 0.520 | 0.951 | 0.605–1.497 | 0.830 |
| Marital status: Single | 1.223 | 0.798–1.873 | 0.360 | 1.292 | 0.835–1.998 | 0.250 |
| Contact: Not provided | 1.105 | 0.781–1.565 | 0.570 | 1.157 | 0.807–1.659 | 0.427 |
| Weight (per kg) | 1.023 | 1.005–1.042 | 0.014 | 1.013 | 0.996–1.031 | 0.144 |
| Hospital admission: No | 0.726 | 0.485–1.085 | 0.120 | 0.703 | 0.458–1.079 | 0.107 |

Long travel distance to the ART facility remained the only statistically significant predictor under both modelling frameworks: Fine-Gray subdistribution hazard = 1.493 (95% CI: 1.079–2.064, *p* = 0.015) and cause-specific HR = 1.454 (95% CI: 1.030–2.053, *p* = 0.033). This consistency in both direction and magnitude indicates that travel distance is a robust determinant of LTFU, influencing both the cumulative incidence of LTFU in the population and the instantaneous risk among individuals who remain event-free.

One noteworthy divergence was observed for baseline weight. Under the Fine-Gray model, higher weight was significantly associated with an increased SHR of LTFU (SHR = 1.023, 95% CI: 1.005– 1.042, *p* = 0.014), whereas the corresponding cause-specific hazard ratio was similar in magnitude but not statistically significant (CS-HR = 1.013, 95% CI: 0.996–1.031, *p* = 0.144). This attenuation is consistent with the interpretation that heavier patients experience a lower competing risk of death (death SHR = 0.932, *p <* 0.001), which influences the cumulative incidence of LTFU while exerting less influence on the cause-specific hazard. Consequently, the Fine-Gray estimate provides the more appropriate population-level summary measure. For all remaining covariates, both models produced non-significant estimates in the same direction, indicating good overall agreement.

#### 3.5.2 Cause-Specific Sensitivity Analysis for Death

Table 6 presents the corresponding comparison for mortality. The cause-specific model identified 52 deaths among 329 patients (concordance *C* = 0.79; likelihood ratio test: *χ*^2^ = 63.1, *df* = 10, *p* = 9 × 10*^−^*^10^). The results were highly consistent with those obtained from the Fine-Gray model across nearly all covariates.

**Table 6:**
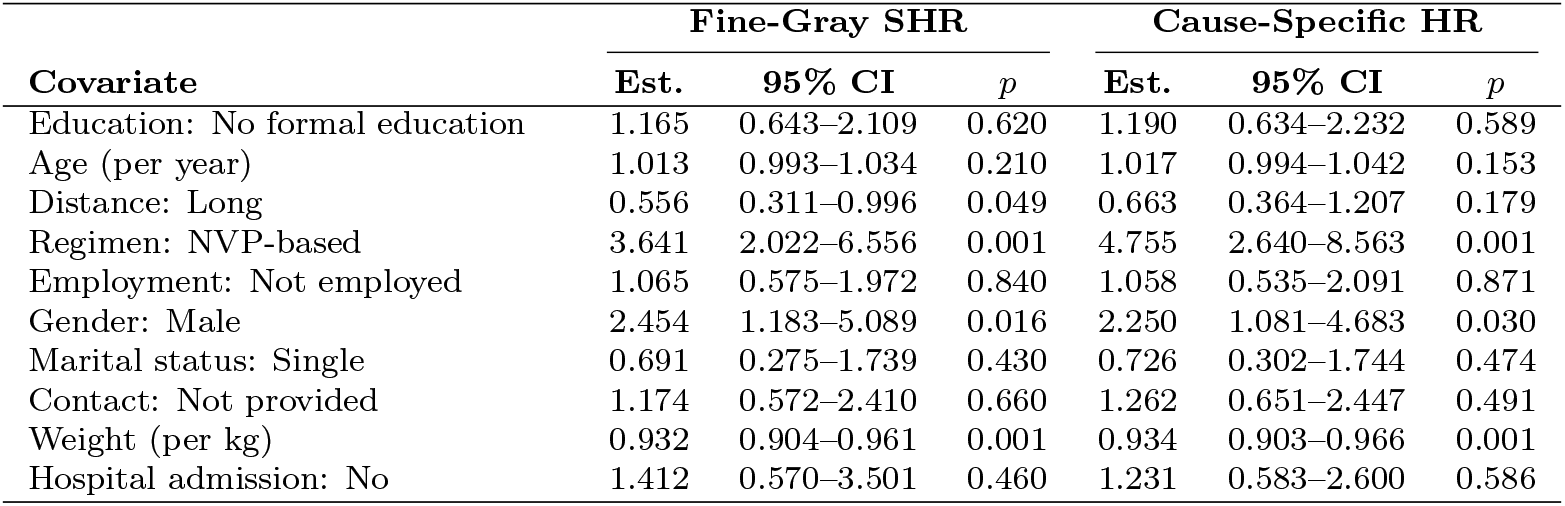
Performance comparison of Fine-Gray SHR and CS-HR for death.

| Covariate | Fine-Gray SHR |  |  | Cause-Specific HR |  |  |
| --- | --- | --- | --- | --- | --- | --- |
| | Est. | 95% CI | $p$ | Est. | 95% CI | $p$ |
| Education: No formal education | 1.165 | 0.643–2.109 | 0.620 | 1.190 | 0.634–2.232 | 0.589 |
| Age (per year) | 1.013 | 0.993–1.034 | 0.210 | 1.017 | 0.994–1.042 | 0.153 |
| Distance: Long | 0.556 | 0.311–0.996 | 0.049 | 0.663 | 0.364–1.207 | 0.179 |
| Regimen: NVP-based | 3.641 | 2.022–6.556 | 0.001 | 4.755 | 2.640–8.563 | 0.001 |
| Employment: Not employed | 1.065 | 0.575–1.972 | 0.840 | 1.058 | 0.535–2.091 | 0.871 |
| Gender: Male | 2.454 | 1.183–5.089 | 0.016 | 2.250 | 1.081–4.683 | 0.030 |
| Marital status: Single | 0.691 | 0.275–1.739 | 0.430 | 0.726 | 0.302–1.744 | 0.474 |
| Contact: Not provided | 1.174 | 0.572–2.410 | 0.660 | 1.262 | 0.651–2.447 | 0.491 |
| Weight (per kg) | 0.932 | 0.904–0.961 | 0.001 | 0.934 | 0.903–0.966 | 0.001 |
| Hospital admission: No | 1.412 | 0.570–3.501 | 0.460 | 1.231 | 0.583–2.600 | 0.586 |

NVP-based ART regimen was the strongest predictor of mortality under both modelling approaches. The Fine-Gray model estimated an SHR of 3.641 (95% CI: 2.022–6.556, *p <* 0.001), while the cause-specific model estimated an HR of 4.755 (95% CI: 2.640–8.563, *p <* 0.001). The larger cause-specific estimate is theoretically expected because patients receiving NVP-based regimens are also susceptible to the competing event of LTFU, which attenuates the cumulative incidence estimate.

Male sex was also significantly associated with higher mortality under both approaches (SHR = 2.454, 95% CI: 1.183–5.089, *p* = 0.016; CS-HR = 2.250, 95% CI: 1.081–4.683, *p* = 0.030). Likewise, higher baseline weight remained strongly protective against mortality under both models (SHR = 0.932, 95% CI: 0.904–0.961, *p <* 0.001; CS-HR = 0.934, 95% CI: 0.903–0.966, *p <* 0.001). The near-identical estimates confirm a robust inverse association between baseline nutritional status and mortality risk.

A single divergence was observed for travel distance. The Fine-Gray model suggested that long travel distance was associated with a lower cumulative incidence of death (SHR = 0.556, 95% CI: 0.311–0.996, *p* = 0.049), whereas the corresponding cause-specific hazard ratio was directionally similar but not statistically significant (CS-HR = 0.663, 95% CI: 0.364–1.207, *p* = 0.179). This finding suggests that the reduced cumulative incidence of death among patients travelling longer distances is partially explained by their increased probability of experiencing the competing event of LTFU before death.

## 4 Discussion and Conclusion

### 4.1 Principal Findings

This study investigated loss to follow-up among HIV patients receiving antiretroviral therapy while accounting for the competing risk of death using Fine-Gray subdistribution hazard regression. The findings indicate that loss to follow-up occurred more frequently than mortality throughout the follow-up period. Only travel distance and baseline weight were significant predictors of LTFU, whereas ART regimen, sex, and baseline weight were significant predictors of mortality. These findings highlight the importance of both geographical and clinical factors in shaping retention and survival outcomes among patients receiving ART.

### 4.2 Cumulative Incidence of LTFU and Mortality

The cumulative incidence of LTFU reached 50.2% by 36 months of follow-up, indicating that attrition remains a critical challenge in the Northern Ghana ART programme. This magnitude is consistent with prior evidence from comparable West African settings [3], although it contrasts with studies reporting higher LTFU rates among male patients specifically [22], a discrepancy we return to below. The overall incidence of death was 15.8%, exceeding rates reported in Uganda (4.5%) and North-Western Tanzania (11.6%) [13, 19]. Despite this elevated mortality, the cumulative incidence of death increased at a slower and relatively constant rate over the observation window, consistent with the mortality-attenuating effects of sustained ART coverage documented in high-income and low-income settings alike [29].

### 4.3 Predictors of Loss to Follow-Up

Two covariates were independently associated with the cumulative incidence of LTFU: travel distance to the ART facility and baseline weight. Patients residing farther from the treatment centre experienced a significantly higher cumulative incidence of loss to follow-up. This finding is consistent with previous studies reporting that geographical barriers impede sustained engagement in HIV care [6]. Long travel distances may impose transportation costs, increased travel time, and missed work opportunities that discourage routine clinic attendance. In rural settings, poor transportation infrastructure and limited access to healthcare facilities may further reduce adherence to scheduled appointments, thereby increasing the likelihood of treatment interruption and eventual loss to follow-up [16, 17]. These findings highlight healthcare accessibility as an important determinant of programme retention. However, the mechanisms underlying this association were not directly measured in the present study and therefore remain speculative.

Higher baseline weight was associated with an increased cumulative incidence of loss to follow-up but a lower cumulative incidence of mortality. The mechanisms underlying these contrasting associations remain uncertain. One possible explanation is that patients with better perceived health and nutritional status may be less likely to maintain regular clinic attendance after ART initiation, while simultaneously experiencing improved survival outcomes. Better health may also facilitate engagement in employment and other socioeconomic activities that compete with routine clinic visits. Conversely, higher baseline weight may reflect better nutritional status and overall physiological reserve, which are associated with improved treatment tolerance, enhanced immunological recovery, and reduced mortality risk. However, these explanations could not be formally evaluated using the available data and should therefore be interpreted cautiously.

### 4.4 Predictors of Mortality

Three covariates were independently associated with the subdistribution hazard of death: sex, baseline weight, and ART regimen type.

Male patients exhibited a significantly higher cumulative incidence of mortality than female patients. This observation is consistent with previous evidence suggesting that men often initiate HIV treatment later in the disease course, utilize healthcare services less frequently, and may experience delayed diagnosis and treatment initiation [1, 19, 22]. These factors can contribute to poorer clinical outcomes and increased mortality among male patients receiving ART. Men may also use preventive and routine health services less, increasing the risk of treatment interruption and late presentation with opportunistic infections [13, 25]. In many settings, women maintain greater contact with health services through reproductive and maternal health programmes, facilitating earlier HIV diagnosis and linkage to ART [3]. Consequently, male patients tend to initiate therapy with greater immunosuppression and more advanced disease, contributing to poorer survival outcomes.

Higher baseline weight was protective against mortality, a finding consistent with the established role of nutritional status and overall physiological reserve as determinants of ART outcomes [13, 22, 25]. Patients with greater body weight at treatment initiation are generally better able to tolerate ART, maintain immunological recovery, and resist opportunistic infections during the early stages of treatment [8, 29]. This observation likely reflects the adverse impact of undernutrition and poor physiological reserve on HIV disease progression and survival among patients receiving ART [25, 29].

Patients receiving efavirenz (EFV)-based regimens experienced a substantially lower cumulative incidence of mortality than those receiving nevirapine (NVP)-based regimens, with an estimated 72% reduction in the subdistribution hazard of death (SHR = 0.28, 95% CI: 0.15–0.50). This finding is consistent with previous studies reporting superior virological and clinical outcomes among patients receiving EFV-based therapy [4, 7, 10, 20]. Several mechanisms may explain this association. Nevirapine has been linked to serious adverse drug reactions, including hepatotoxicity and severe cutaneous hypersensitivity, which can adversely affect treatment continuation and clinical outcomes. In addition, NVP-based regimens were more commonly prescribed during earlier treatment eras and may therefore reflect less favourable clinical circumstances at treatment initiation. Consequently, the observed survival advantage associated with EFV-based regimens likely reflects a combination of differences in drug toxicity, historical prescribing practices, and residual variation in disease severity at ART initiation. In contrast, ART regimen was not significantly associated with the cumulative incidence of loss to follow-up (SHR = 1.17, 95% CI: 0.81–1.68). Although the point estimate suggested a modest increase in the subdistribution hazard of LTFU among patients receiving EFV-based regimens, the wide confidence interval and lack of statistical significance indicate insufficient evidence of an independent association. While differences in drug tolerability and adverse-effect profiles may plausibly influence retention in care, the present findings do not support regimen type as a major determinant of loss to follow-up in this cohort. Furthermore, treatment guidelines and prescribing practices have evolved substantially since the period during which many NVP-based regimens were initiated, which may limit the contemporary relevance of regimen-related differences in retention.

### 4.5 Comparison of Fine-Gray and Cause-Specific Cox Models

The cause-specific Cox models corroborated the substantive conclusions of the primary Fine-Gray analysis. Across both outcomes and all covariates, most substantive conclusions were consistent across both modelling frameworks, although travel distance was statistically significant only in the Fine-Gray mortality model.

Differences in the magnitude of SHRs and cause-specific hazard ratios (CS-HRs) are theoretically expected and reflect the influence of competing events rather than model instability. When the SHR exceeds the corresponding CS-HR, the covariate elevates the cumulative incidence of the primary outcome both by accelerating that event and by simultaneously reducing the competing event, increasing the pool of individuals remaining susceptible over time. Conversely, when the SHR is smaller than the CS-HR — as observed for the NVP-based regimen in the mortality analysis — the covariate also increases the occurrence of the competing event (LTFU), diverting individuals away from the primary outcome and attenuating its cumulative incidence relative to the underlying cause-specific hazard. These patterns are fully consistent with competing-risks theory. Reporting both modelling frameworks simultaneously provides complementary inferential perspectives, distinguishing predictors of cumulative incidence from predictors of the instantaneous hazard rate, and is recommended as good practice in competing-risks analyses [5, 24].

### 4.6 Strengths and Limitations

This study has several strengths. The competing-risks framework enabled simultaneous evaluation of mortality and loss to follow-up while accounting for their interdependence. In addition, model assumptions were formally assessed, and findings were evaluated through sensitivity analyses.

This study was conducted using retrospective data from 329 patients attending a single tertiary hospital in Northern Ghana. Although the cohort provides valuable insights into factors associated with mortality and loss to follow-up among people living with HIV receiving ART, the single-centre design may limit the external validity of the findings. Healthcare access, patient characteristics, treatment practices, and resource availability may differ across regions and healthcare facilities, potentially affecting the applicability of the results to other settings. Consequently, caution is warranted when extrapolating these findings beyond the study population. Future multicentre studies including patients from diverse geographic and healthcare settings are needed to evaluate the consistency and generalisability of the observed associations.

Several clinically important variables, including CD4 cell count, viral load, WHO clinical stage, antiretroviral therapy adherence, tuberculosis co-infection status, and measures of socioeconomic status, were not available in the retrospective records and therefore could not be incorporated into the competing-risks models. Because these factors are known determinants of HIV treatment outcomes, their omission may have introduced residual confounding and could partially explain some of the observed associations. Although the analyses adjusted for available demographic and clinical characteristics, the possibility of unmeasured confounding cannot be excluded. Consequently, residual confounding may persist despite adjustment for available covariates. Future studies incorporating more comprehensive clinical and behavioural data are warranted to validate these findings.

### 4.7 Clinical and Policy Implications

The findings carry several actionable implications for HIV programme management in Northern Ghana and comparable settings. Because travel distance was the strongest independent predictor of loss to follow-up, interventions should prioritise patients residing far from treatment centres through decentralised ART delivery, community-based drug distribution programmes, and transportation-support initiatives; decentralisation of this kind directly reduces the structural travel burden identified as a barrier to retention [6]. Targeted adherence counselling, particularly for patients showing early clinical improvement who may be at risk of disengagement from care for reasons warranting further investigation, could reinforce the importance of sustained treatment and support earlier identification of patients at risk of disengaging from care. The observed mortality differences between EFV- and NVP-based regimens should be interpreted in the context of historical prescribing practices and the observational nature of the study. Furthermore, current HIV treatment guidelines increasingly recommend integrase inhibitor-based regimens, particularly dolutegravir-based therapy, which may limit the direct applicability of these findings to contemporary clinical practice. Demand-side interventions targeting men who face higher mortality and later presentation, including community screening and male-friendly clinic environments, warrant prioritisation. Collectively, these strategies are aligned with Sustainable Development Goal 3.3 and the UNAIDS 95–95–95 targets for ending AIDS as a public health threat by 2030.

### 4.8 Conclusion

This study applied the Fine-Gray subdistribution hazard model to investigate loss to follow-up among HIV patients receiving antiretroviral therapy while accounting for the competing risk of death. The results demonstrated that travel distance to the treatment facility and baseline body weight were significant predictors of the cumulative incidence of loss to follow-up, whereas sex, baseline weight, and ART regimen type were significant predictors of mortality. These findings emphasize the importance of addressing structural barriers to care and strengthening patient-retention strategies within HIV treatment programmes. The study further demonstrates the value of competing-risk methodology for obtaining more accurate estimates of event probabilities and covariate effects in the presence of competing outcomes. Future research incorporating additional clinical and socioeconomic variables from multiple treatment centres may provide a more comprehensive understanding of factors influencing retention and survival among individuals receiving ART.

## 5 Author Contributions

Mohammed Yushawu Abdulai conceived and designed the study, conducted the data collection, and reviewed the drafted manuscript. Solomon Crusader Abo designed the methodology, performed the data analysis and interpretation, drafted the manuscript, and revised it critically for important intellectual content. Benjamin Tommy Bavug contributed to data cleaning, participated in and validated the writing, review, and editing of the manuscript. All authors contributed their individual resources, read and approved the final version of the manuscript, and agree to be accountable for all aspects of the work to ensure that questions related to the accuracy or integrity of any part of the work are appropriately investigated and resolved.

## 8 Acknowledgments

The authors gratefully acknowledge the management, staff, and data management team of the Antiretroviral Therapy (ART) Unit of the Baptist Medical Centre, Nalerigu, for granting access to the study records and supporting the conduct of this research. We also acknowledge the patients whose anonymized records contributed to this analysis.

## 9 Data and Code Availability Statement

This study used patient-level clinical records from the Antiretroviral Therapy (ART) Unit of the Baptist Medical Centre, Nalerigu, Ghana. Public release of these data is restricted in accordance with the ethical approval granted by the Committee on Human Research, Publication and Ethics (CHRPE), School of Medicine and Dentistry, Kwame Nkrumah University of Science and Technology (Approval No. CHRPE/AP/1181/26). A minimal de-identified dataset sufficient to reproduce the findings has been deposited in Zenodo [2] under controlled access. Access to the dataset is restricted because it contains sensitive patient-level information and is subject to institutional and ethical approval requirements. Qualified researchers may request access through the Baptist Medical Centre ART Unit, Nalerigu, Ghana. The data dictionary and all R scripts used for data management, statistical analyses, simulation studies, and figure generation have been deposited in Zenodo [2] and are available at: https://doi.org/10.5281/zenodo.23269219 The analytical code and supplementary simulation code are also available in the GitHub repository: https://github.com/Nanaabo/Fine-Gray-Competing-risks-Sim

## A Assessment of the Proportional Subdistribution Hazard Assumption

Figure 2 and Figure 3 display the Schoenfeld residual plots for the lost-to-follow-up and death subdistribution hazard models, respectively.

**Figure 2:**
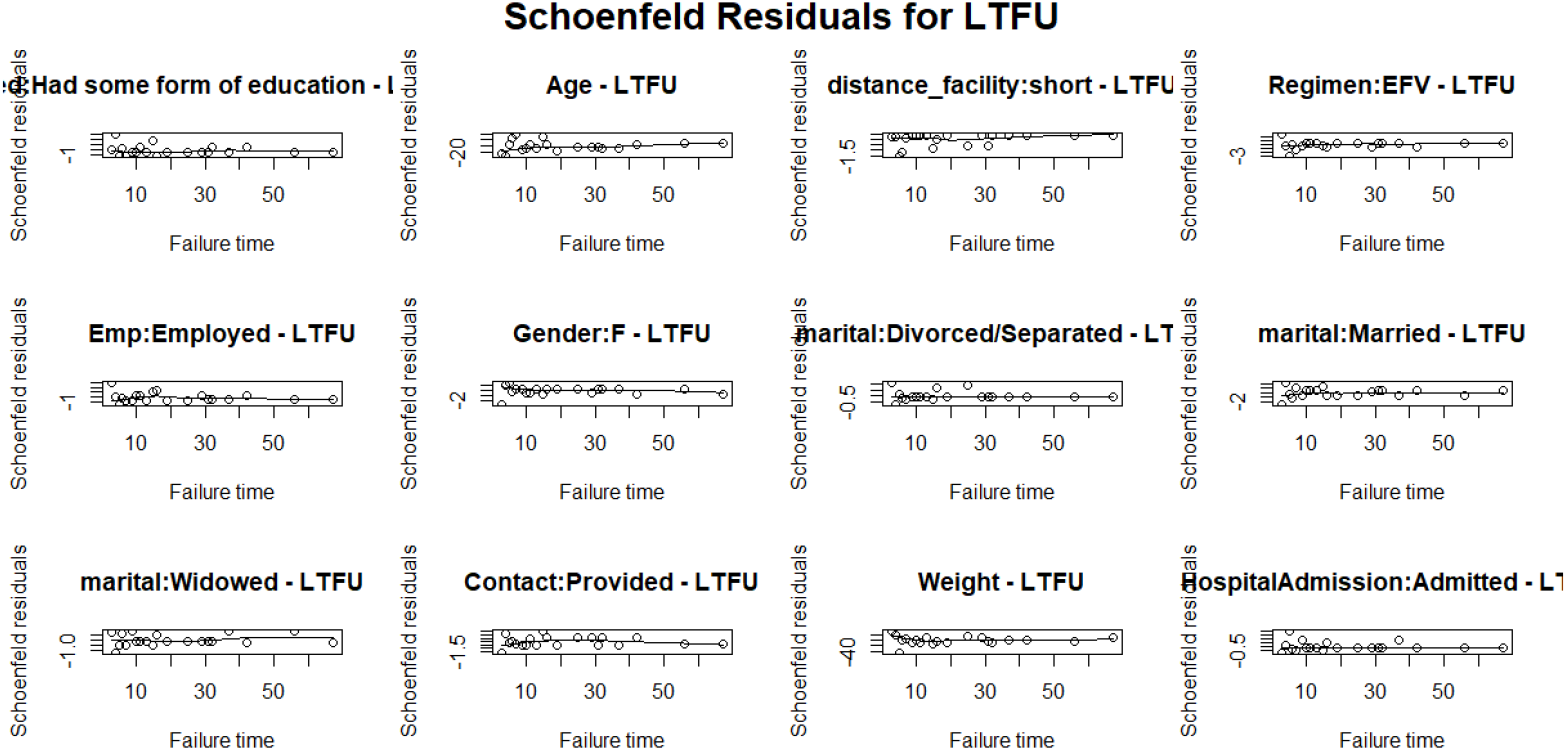
Schoenfeld residuals with Loss to follow-up as the event of interest

**Figure 3:**
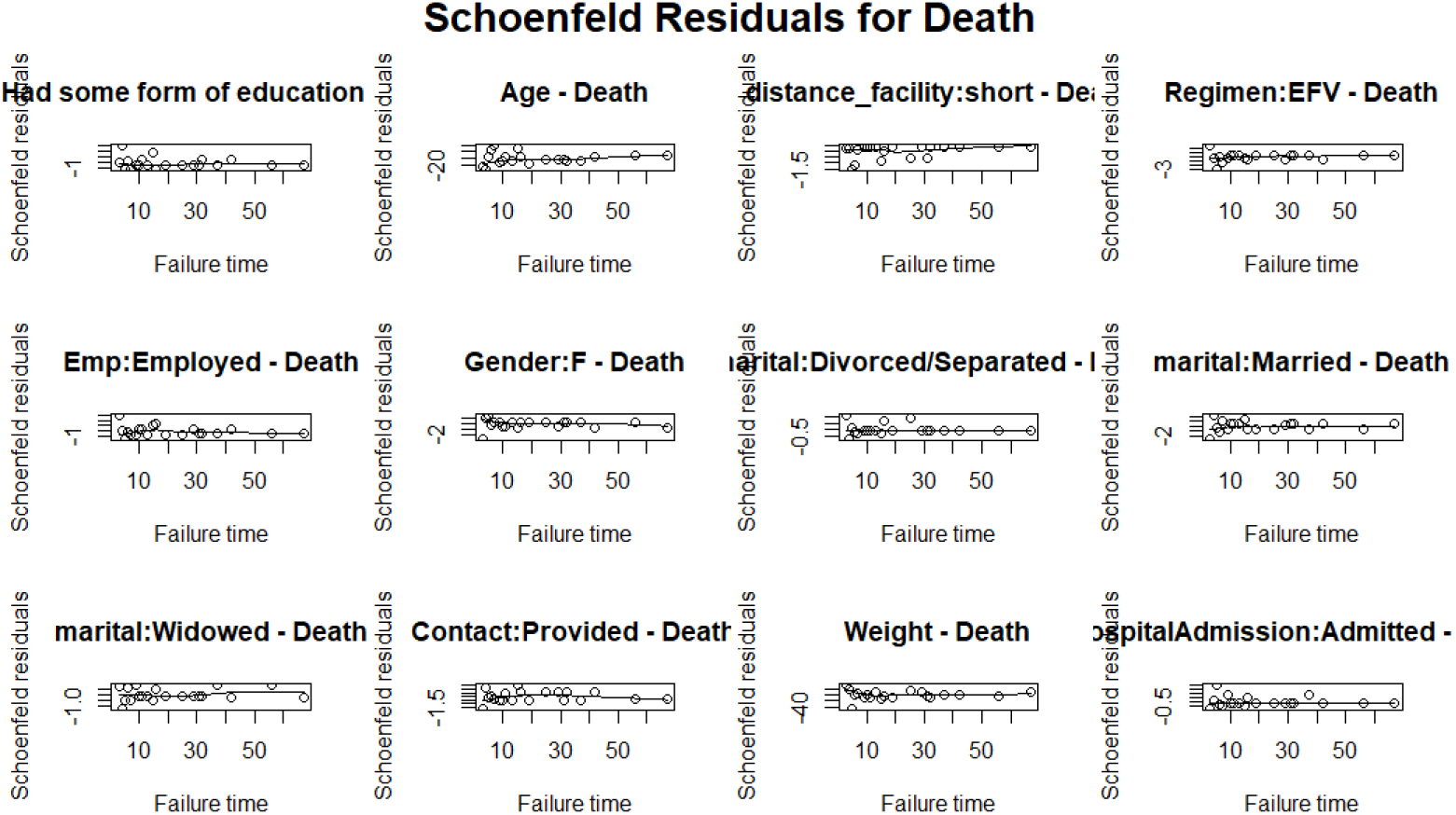
Schoenfeld residuals with Death as the event of interest

## B Socio-Demographic Characteristics of Patients by Gender

**Table A1:** Socio-Demographic Characteristics of Patients by Gender.

| Characteristic | Female<br>($n = 232$) | Male<br>($n = 97$) | Overall<br>($n = 329$) |
| --- | --- | --- | --- |
| *Age group (years)* |  |  |  |
| 15–29 | 110 (47.4) | 6 (6.2) | 116 (35.3) |
| 30–49 | 107 (46.1) | 70 (72.2) | 177 (53.8) |
| 50+ | 15 (6.5) | 21 (21.6) | 36 (10.9) |
| *Religion* |  |  |  |
| Christian | 98 (42.2) | 25 (25.8) | 123 (37.4) |
| Muslim | 121 (52.2) | 55 (56.7) | 176 (53.5) |
| Traditionalist | 13 (5.6) | 17 (17.5) | 30 (9.1) |
| *Marital status* |  |  |  |
| Divorced/Separated | 10 (4.3) | 8 (8.2) | 18 (5.5) |
| Married | 154 (66.4) | 64 (66.0) | 218 (66.3) |
| Single | 37 (15.9) | 19 (19.6) | 56 (17.0) |
| Widowed | 31 (13.4) | 6 (6.2) | 37 (11.2) |
| *Education* |  |  |  |
| No formal education | 137 (59.1) | 49 (50.5) | 186 (56.5) |
| Some formal education | 95 (40.9) | 48 (49.5) | 143 (43.5) |
| *Employment status* |  |  |  |
| Employed | 88 (37.9) | 88 (90.5) | 176 (53.5) |
| Not employed | 144 (62.1) | 9 (9.3) | 153 (46.5) |
| *Distance to facility* |  |  |  |
| Long | 102 (44.0) | 52 (53.6) | 154 (46.8) |
| Short | 130 (56.0) | 45 (46.4) | 175 (53.2) |

*Note:* Values are $n$ (%). Percentages may not sum to 100 due to rounding.

## C Simulation Study

To evaluate the finite-sample performance of the Fine–Gray subdistribution hazard estimator under a competing-risks framework similar to that observed in the study cohort, a Monte Carlo simulation study was conducted [2, 11]. Five sample sizes (*n* ∈ {100, 200, 500, 800, 1,000}) were considered, with *R* = 1,000 replicated datasets generated for each scenario. The simulation assessed parameter recovery, estimator bias, standard errors, root mean squared errors (RMSEs), empirical coverage probabilities of nominal 95% confidence intervals, and the effect of increasing sample size on estimator performance. Overall, parameter estimates exhibited relatively small bias, while precision improved with increasing sample size, as reflected by decreasing standard errors and RMSEs. Coverage probabilities for the loss-to-follow-up and censoring parameters remained close to the nominal 95% level across scenarios. However, coverage probabilities for some mortality-related parameters declined at larger sample sizes, indicating potential finite-sample limitations. Collectively, the findings support the practical application of the Fine–Gray model for competing-risks analyses under data-generating mechanisms comparable to those observed in the present ART cohort. Full details of the simulation design, performance measures, and results are provided in the Supplementary Appendix. The R code used to reproduce the simulation study is publicly available at: https://github.com/Nanaabo/Fine-Gray-Competing-risks-Sim

Two competing failure types were considered: Event 1 (loss to follow-up, LTFU) and Event 2 (death). A single binary covariate representing gender was generated as

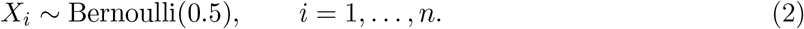

The cause-specific hazards for LTFU and death were specified as

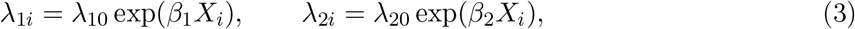

where *λ*_10_ and *λ*_20_ denote baseline hazards and *β*_1_ and *β*_2_ represent the true covariate effects for LTFU and death, respectively. Latent event times were generated independently from exponential distributions:

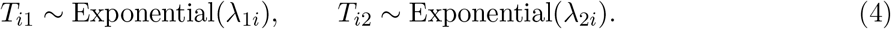

The observed event time was defined by the earliest competing event, *T_i_* = min(*T_i_*_1_*, T_i_*_2_), with event indicator

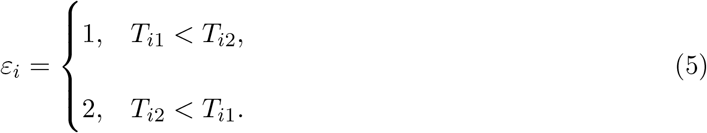

To mimic incomplete follow-up, censoring times were generated independently as *C_i_* ∼ Exponential(*λ_C_*), where *λ_C_* was chosen to yield approximately 20%–30% censoring. The observed follow-up time was defined as 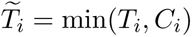, with observed status indicator

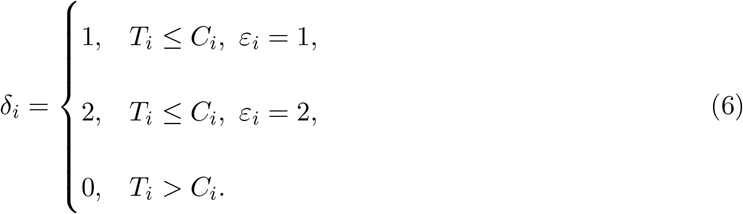

The true parameter values were fixed at *β*_1_ = log(1.20) = 0.1823 and *β*_2_ = log(0.80) = −0.2231, corresponding respectively to a 20% increase in the subdistribution hazard of LTFU and a 20% reduction in the subdistribution hazard of death associated with male gender. The Fine-Gray subdistribution hazard model was fitted separately for each outcome within every replicate, with the other outcome treated as the competing event.

### C.1 Performance Metrics

Let 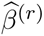 denote the estimated coefficient of parameter *β* in replicate *r*, and let *β^∗^* denote the corresponding true value. Five metrics summarised performance.

#### Average Estimate

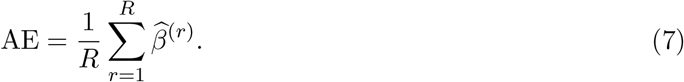

Provides a Monte Carlo estimate of the expected value of the estimator and serves as the basis for computing bias.

#### Bias

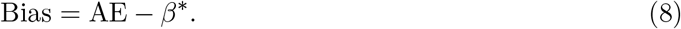

Measures systematic over- or underestimation. Values close to zero indicate negligible systematic error.

#### Standard Deviation (SD)

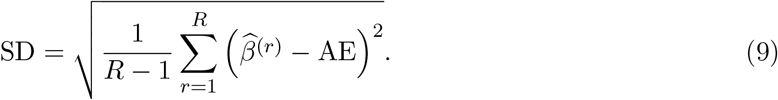

Quantifies Monte Carlo variability of the estimator. Large SD relative to Bias indicates that estimation error is predominantly stochastic rather than systematic.

#### Root Mean Squared Error (RMSE)

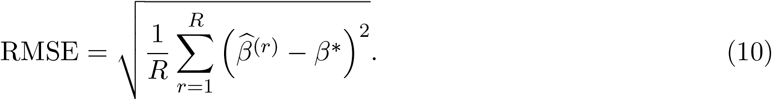

Combines bias and variance into a single accuracy summary via the decomposition RMSE^2^ = Bias^2^ + SD^2^.

#### Coverage Probability (CP)

For each replicate *r*, the nominal 95% confidence interval is

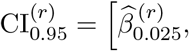

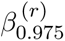, (11) where 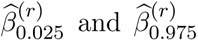 denote the 2.5th and 97.5th percentiles of the confidence limits in replicate *r*. The empirical coverage probability is

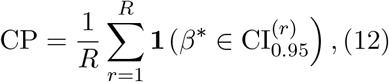

where **1**(·) is the indicator function. Values close to 0.95 indicate well-calibrated intervals; values below the Monte Carlo tolerance band [0.936, 0.964] for *R* = 1,000 replicates indicate under-coverage.

### C.2 Simulation Results

**Table A2:**
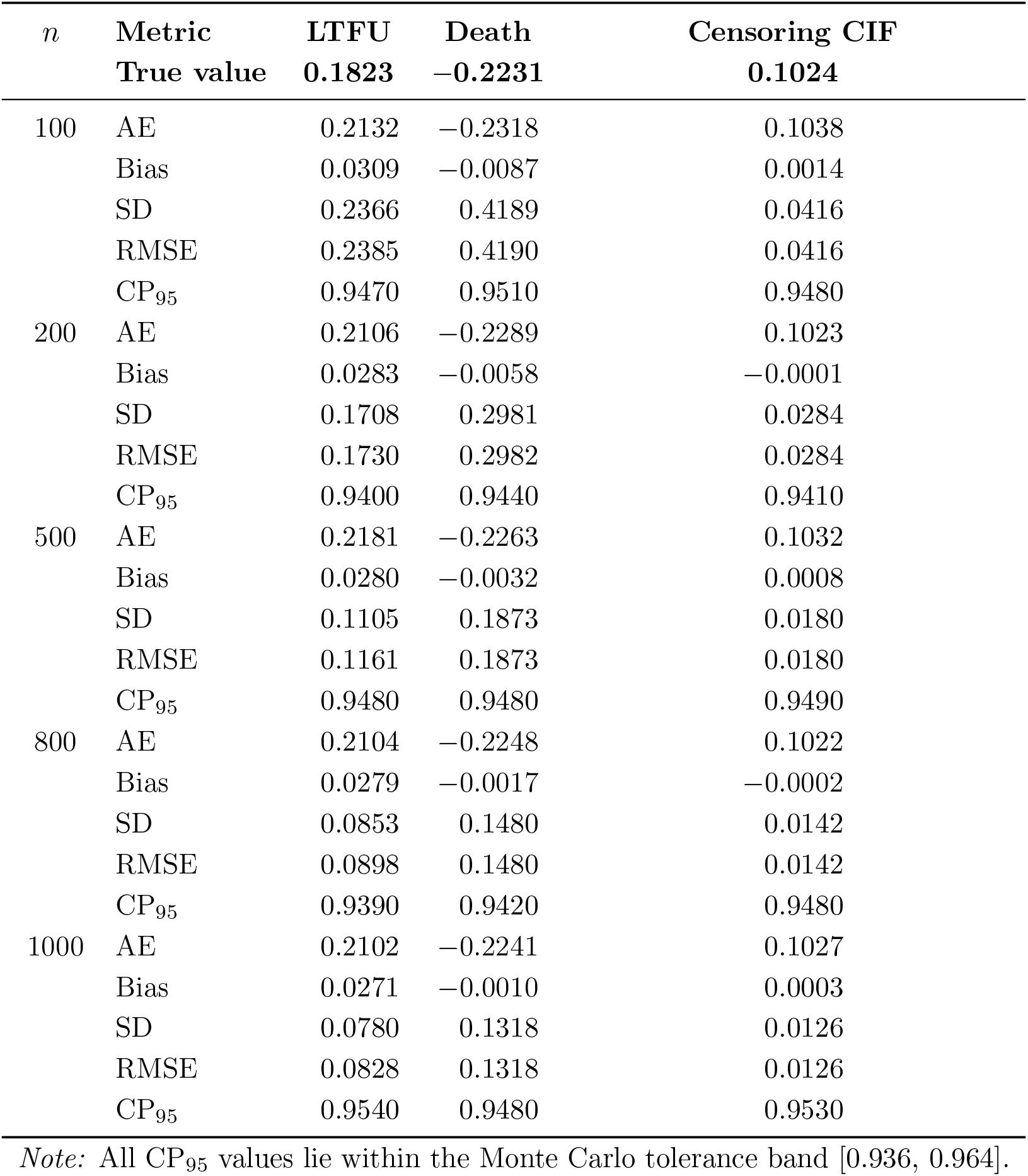
Simulation results for the gender covariate effect on loss to follow-up (LTFU), death, and the censoring cumulative incidence function (CIF) across five sample sizes (*R* = 1,000 replicates). True values: *β*_LTFU_ = 0.1823, *β*_Death_ = −0.2231, *β*_CIF_ = 0.1024. CP_95_ denotes the empirical coverage probability of the nominal 95% confidence interval.

The absolute bias generally decreases as sample size increases, consistent with the theoretical property of estimator consistency under the Fine-Gray model. For the censoring CIF, bias contracts from 0.0014 at *n* = 100 to 0.0003 at *n* = 1,000 (a 79% reduction). The death parameter exhibits the steepest relative reduction, declining from −0.0087 to −0.0010 (89%), confirming that the subdistribution hazard formulation appropriately accommodates the competing-risk structure induced by LTFU. The LTFU parameter shows a consistent reduction from 0.0309 to 0.0271 across the same range; its residual small positive bias is consistent with the known finite-sample upward bias of the subdistribution partial likelihood under moderate censoring and is negligible relative to the corresponding RMSE.

Both SD and RMSE decline substantially with increasing *n* for all three parameters, supporting the *O*(*n^−^*^1^*^/^*^2^) convergence rate expected under the Fine-Gray framework. For the censoring CIF, RMSE reduces from 0.0416 to 0.0126 (threefold improvement); for the death parameter, from 0.4190 to 0.1318 (69% reduction). The near equality of RMSE and SD at all sample sizes — most clearly at *n* = 100 for the death parameter (RMSE = 0.4190, SD = 0.4189) — confirms that residual estimation error is driven by sampling variability rather than systematic bias, consistent with a well-specified estimator.

The empirical 95% coverage probabilities remain within the Monte Carlo tolerance band [0.936, 0.964] across all sample sizes and all three parameters. CP_95_ neither declines nor deteriorates systematically with increasing *n* — the pattern that would indicate model misspecification or incorrect handling of informative censoring. These results collectively support the use of the Fine-Gray subdistribution hazard model as a well-calibrated and consistent inferential framework for competing-risks data in which LTFU constitutes an informative competing event for mortality.

